# Inflammatory Bowel Disease Risk Variants Are Associated with an Increased Risk of Skin Cancer

**DOI:** 10.1101/2021.03.01.21252521

**Authors:** Kelly C. Cushing, Xiaomeng Du, Yanhua Chen, LC Stetson, Annapurna Kuppa, Vincent L. Chen, J Michelle Kahlenberg, Johann E. Gudjonsson, Peter DR Higgins, Elizabeth Speliotes

## Abstract

**Background and Aims:** Inflammatory bowel disease is associated with an increased risk of skin cancer. The aims of this study were to determine whether genomic variants associated with IBD susceptibility are also associated with skin cancer susceptibility and if such risk is augmented by the use of immune-suppressive therapy.

**Methods:** The discovery cohort included participants in the UK Biobank (n=408,381). The validation cohort included participants in the Michigan Genomics Initiative (n=51,405). The primary outcome of interest was skin cancer, sub-grouped into non-melanoma (NMSC) and melanoma skin cancers (MSC). Multivariable logistic regression was performed to identify genomic predictors of skin malignancy. Validated SNPs were evaluated for effect modification by immune-suppressive medication.

**Results:** The discovery cohort included 11,079 cases of NMSC and 2,054 cases of MSC. The validation cohort included 7,334 cases of NMSC and 3,304 cases of MSC. Thirty variants were associated with risk of NMSC in the discovery cohort, of which six replicated in the validation cohort [Increased risk: rs7773324-A (*DUSP22; IRF4*), rs2476601-G (*PTPN22*), rs1847472-C (*BACH2*), rs72810983-A (*CPEB4*); Decreased risk: rs6088765-G (*PROCR; MMP24*), rs11229555-G (*ZFP91-CNTF; GLYAT*)]. Twelve variants were associated with risk of MSC in the discovery cohort, of which three replicated in the validation cohort (Increased risk: rs61839660-T (*IL2RA*); Decreased risk: rs17391694-C (*GIPC2; MGC27382*), rs6088765-G (*PROCR; MMP24*)]. No effect modification was observed.

**Conclusion:** The results of this study highlight shared genetic susceptibility across IBD and skin cancer, with increased risk of NMSC in those who carry risk variants in *IRF4, PTPN22, CPEB4*, and *BACH2* and increased risk of MSC in those who carry a risk variant in *IL2RA*.

## BACKGROUND

Inflammatory bowel disease (IBD) is a chronic immune-mediated disease of the gastrointestinal tract. Patients with IBD are at higher risk for both intestinal and extra-intestinal malignancies.^1^ Traditionally, intestinal malignancies have been attributed to the secondary effects of disease-related chronic inflammation.^2–4^ Alternatively, extra-intestinal malignancies have been attributed to the effects of immune suppressive therapy. However, recent epidemiologic data suggests that there may be an increased risk of extra-intestinal malignancies in IBD, which are mediated via medication-independent effects.^5,6^ This work underscores the importance of further investigation into the pleiotropic effects of IBD-risk variants. In this study, we sought to investigate if genomic variants which increase the risk of IBD also increase the risk of a common extra-intestinal malignancy, skin cancer.

Increased risk of both non-melanoma (NMSC) and melanoma skin cancers (MSC) have been described among patients with IBD.^7^ However, the exact mechanisms by which skin cancers occur in the IBD population remains unclear. NMSC includes both squamous cell and basal cell carcinomas. In 2012, there were estimated to be more than 5 million cases of NMSC in the United States, with a relatively equal breakdown between squamous and basal cell carcinomas.^8^ The incidence rate of NMSC in IBD has been shown to be increased above the general population (IRR 1.64, 95% CI 1.51–1.78).^9^ This increased risk of NSMC has been linked with the use of immune suppressive medications. In a study using a national claims dataset, both recent (OR 3.56, 95% CI 2.81–4.50) and persistent (OR 4.27, 95% CI 3.08–5.92) thiopurine use were associated with increased risk of NMSC in IBD.^9^ The increased risk of NMSC with thiopurine use has been confirmed in several studies including a large prospective observational cohort from France (HR 5.9, 95% CI 2.1–16.4)^10^ as well as a separate national claims dataset (OR 1.85, 95% CI, 1.66–2.05)^7^. In addition, a recent systematic review highlighted that a majority of studies have shown a higher risk of NMSC with thiopurine use.^11^

Significant associations between anti-tumor necrosis factor (anti-TNF) therapies and NMSC have also been observed in several studies across many autoimmune diseases. First, in the nested case-control study referenced above, the authors showed that recent (OR 2.07, 95% CI 1.28–3.33) and persistent (OR 2.18, 95% CI 1.07–4.46) anti-TNF use were associated with an increased risk of NMSC in Crohn’s disease.^9^ Second, in a meta-analysis of six studies with 123,031 individuals, anti-TNF use was associated with an increased risk of NMSC in rheumatoid arthritis (RR 1.28, 95% CI 1.19–1.38).^12^ Third, in a study using data from 13 clinical trials, anti-TNF use was associated with an increased risk of NMSC in psoriasis (SIR 1.76, 95% CI 1.26–2.39).^13^ Therefore, both thiopurines and anti-TNFs are considered to be risk factors for the development of NMSC.

In 2011, there were more than 65,000 cases of MSC in the United States with over 9,000 melanoma-related deaths.^14^ The risk of MSC has been found to be independently associated with IBD, with conflicting evidence regarding the impact of anti-TNF use. The independent association between MSC and IBD was reported in a systematic review and meta-analysis, which utilized 179 cases across 12 studies.^5^ In this manuscript, authors reported an increased risk of MSC in IBD patients compared to the general population (RR 1.37, 95% CI 1.10–1.70). This risk persisted when including only those studies which were completed prior to introduction of anti-TNFs (RR 1.52, 95% CI 1.02–2.25) and was not related to thiopurine use (RR, 1.10, 95% CI 0.73–1.66) suggesting that the increased MSC risk in IBD patients was a medication-independent effect. There has also been data supporting a medication-related increase in risk, specifically with anti-TNFs. In a nested case-control study of claims data from the LifeLink Health Plan, each IBD patient with MSC was matched to 4 IBD patients without MSC. The results of the study showed that anti-TNF therapies were associated with an increased risk of MSC (OR, 1.88; 95% CI, 1.08–3.29).^7^ However, subsequent data has been conflicting.^5,15^

The medication-independent association between MSC and IBD raises the possibility that there may be biologic mechanisms inherent to IBD, which also predispose to skin cancer. Interestingly, it was recently shown that patients with pediatric onset Crohn’s disease are at a higher risk of developing malignancies later in life compared to controls, independent of thiopurine or anti-TNF use.^6^ These data suggest that mechanisms inherent to the pathogenesis of IBD may also be important in cancer pathogenesis. This is not surprising given the critical role of the immune system in both autoimmune disease and tumor surveillance. Furthermore, the skin and the intestine are highly similar organs, both constantly being exposed to environmental pathogens necessitating a level of immune homeostasis (*i*.*e*. a balance between immune tolerance to non-pathogenic exposures and immune activation against pathogenic exposures). Therefore, shared immune perturbations which confer risk of both intestinal inflammation and skin malignancy are biologically plausible.

As the use of immune suppressive therapies continues to expand, a deeper understanding of the impact of IBD risk variants on skin malignancy would help to better direct therapeutic choices. In this study, we aimed to determine 1) if the genetic variants which increase the risk of IBD also increase the risk of skin cancer, 2) if such risk is medication-independent, and 3) if there are gene-medication interactions which confer multiplicative risk.

## METHODS

### Study Design

This study was conducted using a retrospective case-control study design, with two prospectively recruited cohorts (Discovery: UK Biobank; Validation: Michigan Genomics Initiative). All European ancestry cohort participants were eligible for inclusion in the study. Participants were not restricted to a diagnosis of IBD, which would have significantly limited power to detect small to moderate size genomic effects which are common in polygenic diseases and the ability to test the hypotheses posed without such restriction.

### Study Cohorts

#### Study UK Biobank (UKBB)

The UKBB is a prospective population-based cohort of middle aged individuals (40–69 years) who were recruited between the years 2006 and 2010 at over 20 centers to capture ethnic, socioeconomic, and geographic diversity.^16^ Comprehensive data including completion of an electronic consent and self-reported questionnaire, as well as collection of bio-specimens, were obtained at the index visit. The initial ∼50,000 participants were genotyped using the Affymetrix UK BiLEVE Axiom array.^17^ The remainder of participants were genotyped using the Affymetrix UK Biobank Axiom® array. There is a high degree of overlap between the two arrays with over 95% commonality. The analyses in this study were conducted through the UK BioBank Resource Project 18120 (awarded to EKS).

#### Michigan Genomics Initiative (MGI)

The MGI is an ongoing institutional bio-repository (2013-current), which provides approved investigators access to genotype and phenotype data. Informed consent and genotyping was completed through the MGI. DNA was genotyped using the Illumina Human Core Exome Array. Clinical data, including demographics, medical diagnoses, and medication use, were extracted from the electronic medical record. Only individuals of European ancestry were included in this study. A total of 5,496 individuals of non-European ancestry were excluded. This study was conducted with approval from the University of Michigan Institutional Review Board (HUM00159951).

### IBD Risk Variant Selection

First, all established and novel SNPs from the largest meta-analysis of IBD susceptibility to date were included.^18^ This study cohort included 42,992 cases and 53,536 controls. As this study included a diverse population and our analysis was restricted to European only ancestry, we chose only those SNPs which associated with IBD susceptibility in the European cohort (n=232). We concurrently selected SNPs from a recent study in which authors fine mapped IBD susceptibility loci to single variant resolution.^19^ Specifically, the authors identified 18 single causal variants with greater than 95% certainty and 27 variants with greater than 50% certainty (n=45). Therefore, a total of 277 SNPs were selected for inclusion in the study.

Among these 277 SNPs, we identified independent loci using a distance based criteria (>1 mega base). For groups of variants located within 1 mega base of each other, a ranking algorithm was applied to preferentially select the representative SNP from the region. Highest priority group was given to those variants which were mapped to single variant resolution in the fine mapping study. Second priority was given to those SNPs which were identified as lead variants where signals mapped to 2-50 variants, also in the fine mapping study. Final priority was based on the reported p value from the meta-analysis data. A total of 188 variants remained for subsequent analyses.

### Outcomes of Interest and Covariates

#### UKBB

The primary outcome of interest was skin cancer. Skin cancers were grouped into NMSC and MSC. NMSC was defined using the ICD-10 code “C44 Other malignant neoplasms of skin”. MSC was defined using the ICD-10 code “C43 Malignant melanoma of skin”. Additional covariates of interest included age, gender, body mass index (BMI)^20^, and thiopurine/anti-TNF use^7,9,10,21^. BMI was calculated as the mean of all recorded BMIs in the dataset. BMI values of <10 or >100 were excluded from analyses. Medication data was obtained according to UKBB protocol and reflects patients who reported taking the medication at the index visit with a minority having a repeat assessment. Medications of interest included thiopurines (*i*.*e*., 6-mercaptopurine, azathioprine) and anti-TNFs (regardless of indication). Anti-TNFs were limited to the use of adalimumab as no medication data was available for infliximab, certolizumab, or golimumab. There was also no medication data available for newer IBD therapies such as vedolizumab, ustekinumab, and tofacitinib.

#### MGI

The primary outcome of interest was skin cancer. Skin cancers were grouped into NMSC and MSC. NMSC was defined using the ICD-9 code “173.x” and the ICD-10 code “C44.x”. MSC was defined using the ICD-9 code “172.x” and the ICD-10 code “C43.x”. Additional covariates included age, gender, BMI, and medication use. BMI was classified as the most recent measurement at the time of data extraction. BMI values of <10 or >100 were excluded from analyses. Medication data was defined as ever versus never exposed and limited to prescriptions recorded in the institutional medical record. Medications of interest included thiopurines (*i*.*e*., 6-mercaptopurine, azathioprine) and anti-TNFs (infliximab, adalimumab, certolizumab, and/or golimumab).

### Statistical Analyses

#### Identification of Genomic Variants Associated with Skin Cancer

Multivariable logistic regression was performed to evaluate for genomic predictors of NMSC and MSC while controlling for age (years), gender (reference category: female), BMI, thiopurine use, anti-TNF use, and population stratification (genetic principal components 1-10). We used KING software to exclude samples related up to 2^nd^ degree. For NMSC and MSC respectively, we randomly sampled controls (3 times the case number) to make a balanced case to control ratio. All independent SNPs (n=188) were included in the multivariable logistic regression model in the discovery cohort. SNPs which were associated with the outcome of interest, at a p-value of less than or equal to 0.05, were then carried forward into the multivariable logistic regression model in the validation cohort. SNPs which had a p-value of less than 0.05 and a concordant direction of effect across both cohorts were considered to have replicated. The direction of effect was aligned with the Crohn’s disease risk increasing allele.^18^ All analyses were performed using *R*, version 3.6.0.^22^ To confirm our findings, we also completed a meta-analysis including both cohorts (UKBB and MGI) using the program METAL.^23^

#### Effect Modification

SNPs which replicated in the validation cohort were evaluated for effect modification by immune suppressive therapy. We restricted interaction analyses to thiopurines/genotype and NMSC risk given the low absolute number of medication exposures in other subgroups. A multivariable logistic regression model including main effects of SNPs and their interaction terms, controlling for age, gender, and BMI, was completed. Interaction terms with a p-value of less than 0.05 were considered significant. All analyses were again performed using *R*, version 3.6.0.^22^

## RESULTS

### Study Cohorts

The discovery cohort (UKBB) included a total of 408,381 participants. There were 11,079 (2.7%) cases of NMSC and 2,054 (0.5%) cases of MSC. The mean age of the study cohort was 67 (+/− 8) years and the mean BMI was 27.4 (+/− 4.8). There was relatively equal gender breakdown with 46% of participants being male and 54% being female. Thiopurine medications were used by 911 (0.2%) individuals, with 73 (0.7%) exposures in NMSC cases and 838 (0.2%) exposures in controls. (**Table 1**) Comparatively, thiopurine exposure was recorded in 5 (0.2%) cases of MSC and 906 (0.2%) controls. (**Table 2**) Anti-TNF medications were used by 138 (0.03%) individuals, with 10 (0.1%) exposures among NMSC cases and 128 (<0.1%) exposures among controls. Comparatively, anti-TNFs were recorded in 0 cases of MSC and 138 (<0.1%) controls.

**Table 1.** Absolute Rates of Immune Suppressive Medication Use, Stratified by Non-Melanoma Skin Cancer (NMSC) Development, Among Patients in the Discovery Cohort (UKBB) and the Validation Cohort (MGI)

| UKBB | NMSC (n=11,079) | No NMSC (n=397,302) |
| --- | --- | --- |
| Thiopurine | 73 (0.7%) | 838 (0.2%) |
| Anti-TNF | 10 (0.1%) | 128 (0.0%) |
| MGI | NMSC (n=7,334) | No NMSC (n=44,071) |
| Thiopurine | 113 (1.5%) | 668 (1.5%) |
| Anti-TNF | 101 (1.4%) | 661 (1.5%) |

**Table 2.**
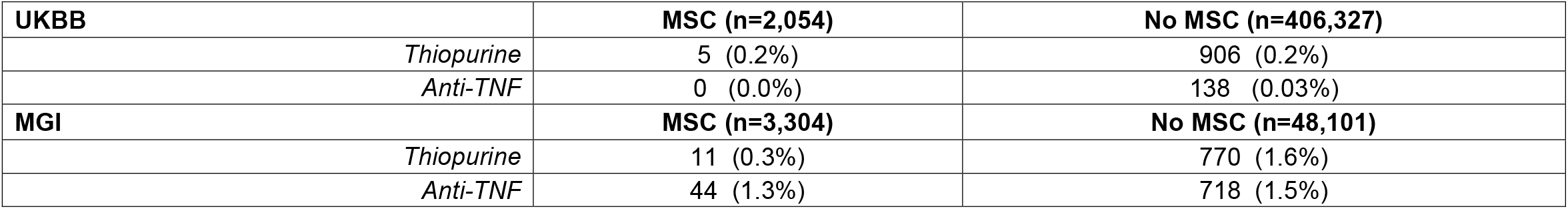
Absolute Rates of Immune Suppressive Medication Use, Stratified by Melanoma Skin Cancer (MSC) Development, in the Discovery Cohort (UKBB) and the Validation Cohort (MGI)

| UKBB | MSC (n=2,054) | No MSC (n=406,327) |
| --- | --- | --- |
| *Thiopurine* | 5 (0.2%) | 906 (0.2%) |
| *Anti-TNF* | 0 (0.0%) | 138 (0.03%) |
| MGI | MSC (n=3,304) | No MSC (n=48,101) |
| *Thiopurine* | 11 (0.3%) | 770 (1.6%) |
| *Anti-TNF* | 44 (1.3%) | 718 (1.5%) |

The validation cohort (MGI) included a total of 51,405 patients. There were 7,334 (14%) cases of NMSC and 3,304 (6.4%) cases of MSC. The mean age was 58 (+/− 16) years, the mean BMI was 30 (+/− 7), and there was again relatively equal gender breakdown (47% male, 53% female). Thiopurine use was recorded in 781 (1.5%) patients, with 113 (1.5%) exposures in NMSC cases and 668 (1.5%) exposures in controls. Comparatively, thiopurine use occurred in 11 (0.3%) cases of MSC and 770 (1.6%) controls. Anti-TNF use was recorded in 762 (1.5%) patients, with 101 (1.4%) exposures among NMSC cases and 661 (1.5%) exposures among controls. Comparatively, anti-TNFs were recorded in 44 (1.3%) cases of MSC and 718 (1.5%) controls.

### Clinical Predictors of Skin Cancer

Age was a significant risk factor for NMSC in the discovery (OR 1.09, 95% CI 1.09,1.1) and validation (OR 1.06, 95% CI 1.06,1.06) cohorts. Age was also a significant risk factor for MSC in the discovery (OR 1.04, 95% CI 1.03,1.05) and validation (OR 1.04, 95% CI 1.03,1.04) cohorts. Male gender was associated with an increased risk of NMSC in both the discovery (OR 1.28, 95% CI 1.22,1.35) and validation (OR 1.27, 95% CI 1.2,1.33) cohorts. However, in MSC, there were paradoxical associations between gender and risk of skin cancer. In the discovery cohort, the risk of MSC was significantly decreased by male gender (OR 0.86, 95% CI 0.77,0.96). However, in the validation cohort, the risk of MSC was significantly increased by male gender (OR 1.38, 95% CI 1.28,1.48). Mean BMI was consistently found to be associated with a decreased risk of NMSC but had no impact on risk of MSC. The impact of BMI on NMSC was small but in the same direction and of similar magnitude across both the discovery (OR 0.98, 95% CI 0.97,0.98) and validation (OR 0.97, 95% CI 0.97,0.98) cohorts. In MSC, BMI was non-significant in both the discovery (OR 0.99, 95% CI 0.99,1.01) and validation (OR 1.00, 95% CI 0.996,1.00) cohorts.

Thiopurine medications were consistently associated with an increased risk of NMSC, similar to previous literature. However, the magnitude of effect was much stronger in the discovery cohort (OR 3.11, 95% CI 2.17,4.45) than the validation cohort (OR 1.38, 95% CI 1.1,1.73). Interestingly, thiopurine use was not associated with risk of MSC in the discovery cohort (OR 1.05, 95% CI 0.38,2.95) but significantly decreased risk in the validation cohort (OR 0.22, 95% CI 0.12,0.40). Anti-TNF therapies were associated with an increased risk of NMSC in the discovery cohort (OR 4.42, 95% CI 1.61,12.1) and were trending towards significance but not significant the validation cohort (OR 1.22, 95% CI 0.97,1.55). The effect of anti-TNFs on MSC in the discovery cohort was not able to be interpreted secondary to a lack of medication exposure events in the outcome of interest. In the validation cohort, anti-TNFs were associated with an increased risk of MSC (OR 1.54, 95% CI 1.12, 2.12). This was an unexpected finding given similar absolute rates on univariate analysis. We therefore performed stratified analyses based on gender, age, and BMI in further investigation. This analysis, included in the supplementary information, revealed paradoxical associations between anti-TNF use and risk of MSC based on age and BMI, with older age and normal BMI conveying higher absolute risk of MSC with anti-TNF exposure. However, multivariable logistic regression revealed no statistically significant interactions between anti-TNF use and age or BMI.

### Genomic IBD Predictors of Skin Cancer

In the discovery cohort, 30 variants associated with the risk of NMSC, all of which were tested for replication in the validation cohort. (**Supplementary Table 1**) Of the 30 variants identified in the discovery cohort, six replicated at a p-value of less than 0.05 with concordant directions of effect: rs7773324-A (*DUSP22; IRF4*: OR 1.14, 95% CI (1.09, 1.19)), rs2476601-G (*PTPN22*: OR 1.14, 95% CI (1.06, 1.21)), rs72810983-A (*CPEB4*: OR 1.07, 95% CI (1.02, 1.11)), rs1847472-C (*BACH2*: OR 1.05, 95% CI (1.01, 1.09)), rs6088765-G (*PROCR; MMP24*: OR 0.95, 95% CI 0.91, 0.99)), and rs11229555-G (*ZFP91-CNTF; GLYAT*: OR 0.95, 95% CI (0.91, 0.99)). (**Table 3**) Variants in *DUSP2/IRF4, PTPN22, CPEB4*, and *BACH2* were associated with an increased risk of NMSC whereas variants in *PROCR/MMP24* and *ZFP91-CNTF/GLYAT* were associated with a decreased risk of NMSC. There were several variants which did not meet statistical significance but had a replicated p-value <0.1 and concordant directions of effect: rs6062496 (p=0.07, *RTEL1-TNFRSF6B*), rs35667974 (p=0.07; Gene: *IFIH1*), and rs727563 (p= 0.09, *ACO2*)

**Table 3.**
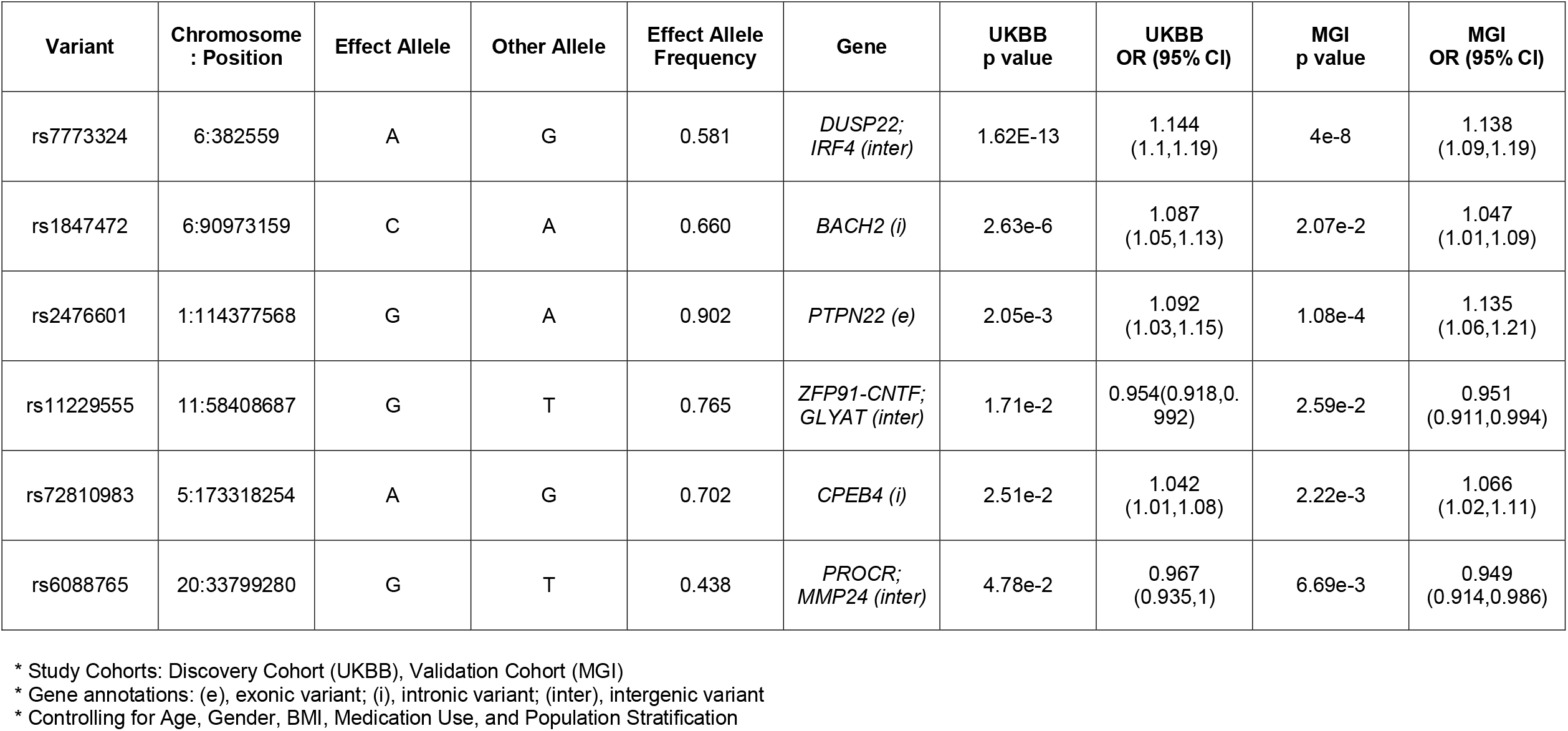
Validated Genomic Predictors of Non-Melanoma Skin Cancer

An additional 12 variants were found to be significantly associated with MSC in the discovery cohort. The 12 identified variants were tested for replication in the validation cohort. (**Supplementary Table 2**) Of the 12 variants in the discovery cohort, three associated with risk of MSC: rs61839660-T (*IL2RA*: OR 1.11, 95% CI (1.02, 1.21)), rs6088765-G (*PROCR; MMP24*: OR 0.92, 95% CI (0.87,0.97)), and rs17391694-C (*GIPC2; MGC27382*: OR 0.92, 95% CI (0.86,0.997)). (**Table 4**) The variant in *IL2RA* associated with an increased risk of MSC whereas variants in *PROCR/MMP24* and *GIPC2/MGC27382* associated with a decreased risk of MSC.

**Table 4.** Validated Genomic Predictors of Melanoma Skin Cancer

| Variant | Chromosome<br>: Position | Effect Allele | Other Allele | Effect Allele<br>Frequency | Gene | UKBB<br>p value | UKBB<br>OR (95% CI) | MGI<br>p value | MGI<br>OR (95% CI) |
| --- | --- | --- | --- | --- | --- | --- | --- | --- | --- |
| rs6088765 | 20:33799280 | G | T | 0.438 | *PROCR*;<br>*MMP24 (inter)* | 1e-3 | 0.871<br>(0.806,0.942) | 1e-3 | 0.918<br>(0.872,0.967) |
| rs61839660 | 10:6094697 | T | C | 0.090 | *IL2RA (i)* | 3e-3 | 1.203<br>(1.06,1.36) | 1.5e-2 | 1.111<br>(1.02,1.21) |
| rs17391694 | 1:78623626 | C | T | 0.879 | *GIPC2*;<br>*MGC27382 (inter)* | 2.2e-2 | 0.885<br>(0.798,0.983) | 4.1e-2 | 0.924<br>(0.857,0.997) |

\* Study Cohorts: Discovery Cohort (UKB B), Validation
Cohort (MGI) \* Gene annotations: (e), exonic variant; (i), intronic variant; (inter), intergenic variant \* Controlling for Age, Gender, BMI, Medication Use, and Population Stratification

In the meta-analysis, all significant variants remained significant. For NMSC, rs7773324 (*DUSP22; IRF4*, p-value 1.12e-18), rs2476601 (*PTPN22*, p-value 1.87e-07), rs72810983 (*CPEB4*, p-value=1.2e-04), rs1847472 (*BACH2*, p-value=1.32e-05), rs6088765 (*PROCR; MMP24*, p-value=1.7e-04), and rs11229555 (*ZFP91-CNTF; GLYAT*, p-value=9.7e-03) were all significant. (**Supplementary Table 3**) For MSC, rs61839660 (*IL2RA*, p-value 6e-04), rs6088765 (*PROCR; MMP24*, p-value 1.88e-06), and rs17391694 (*GIPC2; MGC27382*, p-value 2e-03) were again significant. (**Supplementary Table 4**)

### Gene-Medication Interactions

We next evaluated for interaction effects between immune suppressive medication use and replicated variants. We restricted analyses to NMSC risk and thiopurine-genotype interactions given the small sample sizes in all other groups. The variant rs6088765 was the only genotype which interacted with thiopurines to alter the risk of NMSC in the discovery cohort (p=1.2e-02). (**Table 5**) However, this finding was not replicated in the validation cohort (p=4.2e-01). Therefore, there were no reproducible genotype-thiopurine interactions were identified.

**Table 5.** Genotype-Thiopurine Interactions Effect and Significance in the Risk of NMSC

| **Interaction Term** | **UKBB**<br>**p value** | **UKBB**<br>**OR** | **UKBB**<br>**95% CI** | **MGI**<br>**p value** | **MGI**<br>**OR** | **MGI**<br>**95% CI** |
| --- | --- | --- | --- | --- | --- | --- |
| rs72810983-A | 0.954 | 1.018 | (0.556,1.87) | 0.410 | 1.153 | (0.821,1.62) |
| rs11229555-G | 0.287 | 1.349 | (0.778,2.34) | 0.895 | 0.975 | (0.669,1.42) |
| rs7773324-A | 0.161 | 0.674 | (0.389,1.17) | 0.253 | 1.252 | (0.852,1.84) |
| rs6088765-G | 0.012 | 0.493 | (0.284,0.856) | 0.421 | 1.135 | (0.833,1.55) |
| rs2476601-G | 0.473 | 1.325 | (0.615,2.85) | 0.013 | 0.548 | (0.341,0.881) |
| rs1847472-C | 0.689 | 1.118 | (0.647,1.93) | 0.443 | 0.882 | (0.64,1.22) |

\* Interaction terms refer to the odds ratio associated with thiopurine use multiplied by dosage of the risk allele

## DISCUSSION

Epidemiological studies have demonstrated an increased risk of skin cancer with immune suppressive therapies.^7,9,10^ However, the risk of melanoma has also been associated with IBD independent of medication use suggesting that the relationship between IBD and skin cancer may be more complex than previously thought.^5^ In this study, we sought to determine whether IBD and skin cancer have shared genetic determinants of susceptibility. To answer this question, we performed a retrospective case-control candidate gene study testing known IBD susceptibility variants in two well established genomic biobanks with linkable data on skin cancer diagnoses. The results of this study highlight six IBD susceptibility SNPs which associate with the risk of IBD and NMSC as well as three susceptibility variants which associate with the risk of IBD and MSC.

The variants which associated with the risk of NMSC annotated to the genes *DUSP22/IRF4, PTPN22, CPEB4, BACH2, PROCR/MMP24*, and *ZFP91-CNTF/GLYAT*. Of the identified variants, the most significant association was observed with rs7773324-A, which increased the risk of NMSC by 14%. The rs7773324 locus is located between *DUSP22* and *IRF4*, with closer proximity to the latter. *IRF4* encodes interferon regulatory factor 4, which is a transcription factor expressed in a myriad of immune cells and involved in regulating interferon responses to viral infections.^24^ Interestingly, rs12203592 (T) in *IRF4* has been identified on multiple genome wide association studies to be a strong predictor of squamous cell carcinoma. The first genome wide association study was published in 2016 and used the Kaiser Permanente cohort. In this study, the T allele of rs12203592 was associated with a 56% increased risk of squamous cell carcinoma (OR 1.56, 95% CI 1.49-1.62).^25^ The second GWAS, utilizing cohorts from 23andme as well as the Nurses’ Health Study/ Health Professionals Follow-Up Study, replicated this finding at a similar magnitude of effect (OR 1.62; p-value 2.9×10^−111^).^26^ The *IRF4* rs12203592 T allele has also been found to be associated with an increased risk of invasive squamous cell carcinoma (OR 1.71, 95% CI 1.63-1.80), compared to in situ squamous cell carcinoma, highlighting the role of this gene not only in disease susceptibility but also disease behavior ^27^.

The functional effects of IRF4 in the skin and intestinal tissue have been investigated in previous studies. In the intestine, IRF4 expression is increased in patients with IBD and mediates T_h_17-dependent intestinal inflammation.^28^ In epidermal skin, the intronic *IRF4* variant rs12203592 was found to influence gene expression through interaction with the promoter via a chromatin loop.^29^ The T allele, which is associated with NMSC risk, led to decreased expression of IRF4. The variable expression of IRF across tissue and disease type may be a reflection of the diverse roles of IRF4 in the immune response and would benefit from further investigation.

Interestingly, *BACH2* was also found to associate with NMSC. BACH2 is a transcription factor, which is located in T and B cells^30^ and can directly influence the binding of IRF4 to DNA^31^. BACH2 has an important role in maintaining T cell homeostasis by preventing T cell subset differentiation and promoting T regulatory cell expansion.^30,31^ In our study, the *BACH2* variant (effect allele: C) increased the risk of NMSC by 5-9%. Comparatively, the C allele increases risk of Crohn’s disease by approximately 11%. ^18^ The rs1847472 locus in BACH2 has also been linked with risk of post-operative Crohn’s disease recurrence (OR 1.54, p-value 1.00-2.36). ^32^ The findings of both *IRF4* and *BACH2* as risk loci in IBD and NMSC highlight the importance of further investigation into this signaling network, which may represent an important target for treatment development.

The variant rs2476601-G, located in the gene *PTPN22*, was also associated with an increased risk of NMSC. *PTPN22* encodes protein tyrosine phosphatase non-receptor type 22, which regulates T cell activation.^33^ The missense mutation, rs2476601 (1858 C>T), substitutes a tryptophan for arginine in the first proline-rich C terminal domain.^33^ For ease of reading/reference to our findings, the corresponding base pairs of this mutation will be used going forward [i.e, G>A]. The rs2476601 (A) allele is associated with an increased risk of several autoimmune diseases including but not limited to rheumatoid arthritis, type 1 diabetes mellitus, and psoriatic arthritis.^33^ Paradoxically, the rs2476601 (A) allele is associated with a reduced risk of Crohn’s disease (OR 0.81, 95% CI 0.75–0.89).^34^ The reason behind these discrepant effects remains unclear. In our study, we found that the G allele of rs2476601, which is associated with increased risk of Crohn’s disease ^18^, also increased the risk of NMSC. We found no literature describing an association between *PTPN22* and NMSC; therefore, this is a novel finding.

The rs72810983 A allele, located in *CPEB4* (Cytoplasmic Polyadenylation Element-Binding Protein 4), was also associated with an increased risk of NMSC. *CPEB4* is involved in adenylation/de-adenylation of the poly A tail and has been implicated in the risk of numerous cancer phenotypes including colorectal cancer^35^ and melanoma^36^ as well in microbial diversity of the intestine^37^. Interestingly, we did not see an association between *CPEB4* genotype and MSC risk in our study.

In MSC, the rs61839660 variant (effect allele: T) in *IL2RA* was found to increase risk by 11-20%. The *IL2RA* gene encodes for the alpha subunit of the interleukin-2 receptor, which mediates IL-2 signaling. IL-2 is responsible for T cell homeostasis with effects on both regulatory and effector T cell populations.^38^ Interestingly, high dose IL-2 therapy was found to have anti-tumor effects in animal models^39^, which led to clinical trials studying the efficacy of this therapy for melanoma. Of 270 patients exposed to high dose IL-2 therapy, response was observed in 43 patients (16%, 95% CI 12 to 21%).^40^ High dose IL-2 therapy was therefore approved for the treatment of melanoma in the 1990s but its use has been limited by side effects as well as newer, more effective immunotherapies.^41^ To our knowledge, a genomic association between IL-2 receptor genes and MSC has yet to be demonstrated however. In a recent meta-analysis with 30,134 cases of melanoma and 375,188 controls, association between melanoma and *IL2RA* was not described.^42^ Given the established role of IL-2 in the treatment of melanoma, the finding in this study warrants further investigation. In IBD, rs61839660 (T) is associated with an increased risk of Crohn’s disease (OR 1.28; posterior probability 0.999) but not ulcerative colitis.^19^

Of the six Crohn’s disease risk increasing alleles associated with NMSC, two were associated with a reduced risk of NMSC: rs6088765 (*PROCR; MMP24*) and rs11229555 (*ZFP91-CNTF; GLYAT)*. Similarly, of the three Crohn’s disease risk increasing alleles associated with MSC, two were associated with a reduced risk of MSC: rs6088765 (*PROCR; MMP24*) and rs17391694 (*GIPC2; MGC27382*). The rs6088765 variant (effect allele: G), which decreased risk of both MSC and NMSC, is located between the genes *PROCR* and *MMP24*. The G allele is associated with a small increase in risk of Crohn’s disease (<1%).^18^ An association between MSC and *PROCR* (p=7.43 × 10^−10^) has been demonstrated using expression quantitative trait (eQTL) based analyses in melanocytes.^42^ In addition, eQTL analyses with other skin datasets (skin un-exposed, skin not sun-exposed, and transformed skin fibroblasts) highlighted associations with both *PROCR* (not sun-exposed: p=9.02 × 10^−21^) and *MMP24* (not sun-exposed: p=2.82 × 10-7; sun-exposed: 2.31 × 10^−15^).^42^ *PROCR* is also referred to as *EPCR* or endothelial cell protein C receptor. Endothelial cell protein receptor C is expressed on keratinocytes^43^, innate immune cells^44^, and intestinal epithelial cells^45^. The substrate of endothelial cell protein receptor C is activated protein C, which has anti-inflammatory and wound healing effects.^43^ In active IBD, expression of the endothelial cell protein receptor C has been shown to be significantly reduced on epithelial cells compared to controls.^45^ Thus, the endothelial cell protein receptor C appears to play an important anti-inflammatory role in both the skin and intestine.

Congruent with prior literature, thiopurine medications increased the risk of NMSC in both cohorts and should be considered a risk factor for NMSC. Interestingly, thiopurines displayed no association with MSC in the discovery cohort but a protective effect in the validation cohort. The reasons behind the protective association in the validation cohort remain unclear but, as this effect was not seen consistently across both cohorts, no definitive conclusion can be drawn. Anti-TNFs were associated with an increased risk of NMSC in the discovery cohort but not the validation cohort. This reason behind this difference is likely related to differential sample sizes, with an underpowered UKBB cohort including only 10 cases of NMSC that were anti-TNF exposed. Finally, anti-TNF therapy was associated with an increased risk of MSC in the validation cohort consistent with previous literature.

The strengths of this study include the use of well-established genomic biobanks, the ability to control for medication confounders including thiopurines and anti-TNF therapies, and the replication of findings in an independent cohort. Furthermore, there is an important scientific and clinical novelty to these results. Specifically, these data suggest that there are shared genetic underpinnings between IBD and skin cancer. Extra-intestinal manifestations such as pyoderma gangrenosum, erythema nodosum, primary sclerosing cholangitis, etc are classically described and considered to be linked to the underlying process of inflammatory bowel disease. Comparatively, malignancies are considered to be a separate process influenced by long-standing intestinal inflammation or exposure to immune suppressive therapies. While the medication-skin cancer association remains a clinically important and consistently validated finding, our data also suggest that there may be a shared genetic architecture across IBD and skin cancer. As we look to a future in precision medicine, these data offer the first genomic biomarkers which highlight increased risk of both IBD and skin cancer. Furthermore, this work stresses the importance of further investigation into the pleiotropic effects of IBD risk variants across other disease states including malignancies.

There are several limitations of this study. First, we did not limit individuals studied to only those with a diagnosis of IBD. If we had chosen to implement such a selection strategy, the power to detect small to moderate sized effects, which are common in polygenic diseases, would have been significantly diminished. Furthermore, the questions posed did not depend on a diagnosis of IBD; rather, we sought to test if established IBD variants increase the risk of skin cancer at all and if those effects were modified by medication use. However, it will be beneficial to replicate these findings in a well-powered IBD cohort to determine whether these genetic variants associate with skin cancer risk among IBD patients as well as if these genetic variants alter the overall disease onset and course. Second, participants were restricted to those of European ancestry only. This strategy was specifically chosen due to variation in allele frequencies across different ethnicities, which may confound interpretation of findings. Additional studies in non-European cohorts are needed to determine if these variants predict risk across ethnicity or are European specific. Second, the validation cohort used institutional level data. Therefore, any prescriptions which were ordered outside of the electronic medical record would not have been captured in this study. Third, the medications examined were restricted to thiopurines and anti-TNFs due to lack of data on newer biologic therapies in the discovery cohort as well as small sample sizes in the validation cohort. Further studies controlling for anti-IL-12/23, anti-integrin, and JAK inhibitor therapy will be beneficial in understanding the independence of these risk variants. Finally, comprehensive investigation into gene-medication interactions were restricted due small absolute numbers of medication exposed patients. Therefore, no definitive conclusion can be drawn about genotype-medication interactions.

In summary, we highlight shared genetic risk variants across IBD and skin cancer. Our work stresses the importance of understanding the pleotropic effects of IBD susceptibility SNPs on other diseases including malignancies.

## Data Availability

All results are included in the manuscript.

## ACKNOWLEDGEMENTS

The authors acknowledge the Michigan Genomics Initiative participants, Precision Health at the University of Michigan, the University of Michigan Medical School Central Biorepository, the University of Michigan Advanced Genomics Core, and the Data Office for Clinical and Translational Research for providing data and specimen storage, management, processing, and distribution services, and the Center for Statistical Genetics in the Department of Biostatistics at the School of Public Health for genotype data curation, imputation, and management in support of the research reported in this publication.

