## Supplementary Information for "Inflammatory Bowel Disease Risk Variants Are Associated with an Increased Risk of Skin Cancer"

Stratified gender analyses revealed similar rates of MSC by anti-TNF exposure across females and males. In the female subgroup, 19/1397 (1.4%) of patients with MSC were exposed to anti-TNFs compared to 396/25718 (1.5%) of patients without MSC who were exposed to anti-TNFs. In the male subgroup, 25/1907 (1.3%) of patients with MSC were exposed to anti-TNFs compared to 322/22383 (1.4%) of patients without MSC who were exposed to anti-TNFs. However, analyses stratified on age and BMI did reveal paradoxical associations between anti-TNF use and risk of MSC. For age, older age was defined as greater than or equal to 50 years and younger age was defined as less than 50 years. In the younger age subgroup, 5/419 (1.2%) of patients with MSC were exposed to anti-TNFs compared to 335/15065 (2.2%) of patients without MSC who were exposed to anti-TNFs. In the older age subgroup, 39/2885 (1.4%) of patients with MSC were exposed to anti-TNFs compared to 383/33036 (1.2%) of patients without MSC who were exposed to anti-TNFs. For BMI, normal weight was defined as BMI less than 25 and overweight was defined as BMI greater than or equal to 25. For those individuals with a normal BMI, 14/761 (1.8%) of patients with MSC were exposed to anti-TNFs compared to 193/12355 (1.6%) of patients without MSC who were exposed to anti-TNFs. For individuals in the overweight group, 30/2543 (1.2%) of those who developed MSC were exposed to anti-TNFs compared to 525/35746 (1.5%) without MSC who were exposed to anti-TNFs.

We also completed multivariable logistic regression to assess for significant interactions between anti-TNF use and age/gender/BMI. The main effects and interaction terms were included in the model. The interaction terms age * anti-TNF use (p=0.83), gender * anti-TNF use (p=0.45), and BMI * anti-TNF use (p=0.32) revealed no statistically significant interactions.
